# Clinical Validity of IntelliSpace Cognition Digital Assessment Platform in Mild Cognitive Impairment

**DOI:** 10.1101/2023.02.28.22283846

**Authors:** Willem Huijbers, Gijs van Elswijk, Mandy Spaltman, Mike Cornelis, Ben Schmand, Baraa Alnaji, Maxwell Yargeau, Sarah Harlock, Ryu P. Dorn, Bela Ajtai, Erica S. Westphal, Nandor Pinter

## Abstract

We evaluated a digital cognitive assessment platform, Philips IntelliSpace Cognition, in a cross-sectional cohort of patients diagnosed with mild cognitive impairment (MCI). Performance on individual neuropsychological tests, cognitive domain scores, and Alzheimer’s disease (AD) specific composite scores in MCI were compared with a cohort of cognitively normal adults (CN). The cohorts were matched for age, sex, and education. The performance on all but two neuropsychological tests was worse in the MCI group. After ranking the cognitive domains by effect size, we found that the memory domain was most impaired, followed by executive functioning. The Early AD/MCI Alzheimer’s Cognitive Composite (EMACC) and Preclinical Alzheimer’s Cognitive Composite (PACC) scores were constructed from the digital tests on Philips IntelliSpace Cognition. Both AD-specific composite scores showed greater sensitivity and specificity than the Mini-Mental State Examination, as well as individual neuropsychological tests and individual cognitive domain scores. Together, these results demonstrate the diagnostic value of Philips IntelliSpace Cognition in patients with MCI.

## 1 Introduction

Digital cognitive assessments are increasingly used in patients with mild cognitive impairment (MCI) and Alzheimer’s disease (AD) [1]. The transition from traditional paper-and-pencil tests toward digital assessments is driven by a shortage of qualified neuropsychological staff and an increased demand from an aging population. Digital assessments also provide new opportunities to increase efficiency through automatic analysis, reduce human scoring errors, and quantify behavior in novel ways [2]. Akin to established paper-and-pencil tests, here we validate digitized cognitive tests in a clinical setting.

We evaluated a digital assessment platform, Philips IntelliSpace Cognition, in a cross-sectional cohort of MCI patients. First, we assessed the performance of individual neuropsychological metrics from our digital cognitive battery and compared results with a cohort of cognitively normal older adults (CN) from a previous study that demonstrated that the majority of these digitally assessed metrics are equivalent to their paper-and-pencil counterparts [3]. Second, we evaluated a set of cognitive domain scores derived from a disease-agnostic factor model [4]. This model was established in a larger healthy cohort and the cognitive domain scores derived have not yet been validated in patients with MCI. We expect to replicate findings of studies utilizing non-digital tools that found greater impairments in the memory domain and the executive functioning domain, while other cognitive domains, like visual spatial processing, are likely impaired to a lesser degree [5, 6]. Third, we identified two composite scores specific to Alzheimer’s disease that can be constructed from a combination of outcome measures provided by the digital cognitive battery [7, 8]. Disease-specific composites are used in clinical trials and often estimated by paper-and-pencil based tests. We have constructed the Early AD/MCI Alzheimer’s Cognitive Composite (EMACC) [9] and the Preclinical Alzheimer’s Cognitive Composite (PACC) [10, 11], from the digitally administered tests. Fourth, we compared the sensitivity and specificity of individual neuropsychological tests, the cognitive domain scores, and AD-specific composites.

## 2 Methods

### 2.1 Cohort

83 patients with a clinical diagnosis of mild cognitive impairment (MCI) from an outpatient neurology clinic were included (the Dent Neurologic Institute, Amherst, NY, USA). All 83 patients completed the cognitive assessment after signing an informed consent. This study was approved by the Western Institutional Review Board Copernicus Group and the internal committee for biomedical experiments of Philips. The study is registered at clinicaltrials.gov (NCT04243642).

The study inclusion criteria were based on Alzheimer’s Disease Neuroimaging Initiative criteria [12]. Patients aged 50-90 years who were diagnosed with MCI or amnestic MCI as part of their regular clinical care and a confirmation of those diagnoses within the last 12 months were included. Patients were excluded if they had co-morbid neurological or psychiatric disorders known to affect cognition, vision impairment, or hearing loss not corrected to normal, if they were currently admitted to a hospital, assisted living, nursing home or psychiatric facility, or had a neuropsychological assessment in the past month.

Patients were also excluded from the study if they had any of the following in their medical history: unconsciousness for more than 20 minutes related to traumatic brain injury or a head injury that resulted in an overnight hospital stay, any medical event requiring resuscitation in which they were unresponsive for more than 15 minutes, stroke, chemotherapy treatment within the past two months, current diagnosis or history of substance use or dependence, long-term alcohol abuse or daily alcohol consumption of more than four units, medical marijuana use or recreational marijuana use at least once per week, medications that may affect cognitive test performance (e.g., anticonvulsants, antipsychotics, benzodiazepines, opioids, tricyclic antidepressants, oxybutynin) and sleep apnea with an apnea-hypopnea index equal to or greater than 15. Patient eligibility was confirmed through existing medical records, and the most recent progress notes within 12 months of recruitment and again during the study visit.

As part of the screening process, clinical magnetic resonance imaging (MRI) scans were reviewed. Only patients with brain MRI not older than five years were included. MRI exams were acquired on four different scanners, at 1.5T or 3T field strength, with a mix of 2D and 3D acquisitions. The scan parameters varied between scanners, but each protocol included T1 and T2-weighted, fluid attenuated inversion recovery (FLAIR), diffusion weighted imaging (DWI), as well as susceptibility weighted imaging (SWI). MRI studies were reviewed by a neuroradiologist. Patients with major structural abnormalities (e.g., brain tumor, encephalomalacia, lacunar infarct, hemorrhage, frontotemporal lobar degeneration, acute stroke, cerebral amyloid angiopathy) were excluded. White matter hyperintensities were evaluated on T2-weighted and FLAIR sequences and rated according to Fazekas scoring. Patients with Fazekas score 0, 1 and 2 were included, while patients with score 3 were excluded.

The control group consisted of 83 adults from a cognitively normal U.S. reference population (see statistical methods for details) [3]. The total U.S. reference cohort consists of 687 healthy participants, representative of the U.S. census with regard to age, sex, education, and ethnicity/race. These data were collected in two separate studies between 2019 and 2021 (NCT0380138 / NCT04729257) in four states across the U.S., including New York, Pennsylvania, Florida, and California. We ensured that the cohort only included cognitively normal (CN) older adults, applying the Alzheimer’s Disease Neuroimaging Initiative (ADNI) criteria for healthy controls [12].The group was matched to the MCI sample for age, sex, and education (see statistical methods for details).

### 2.2 Cognitive battery

The cognitive battery was administered in-person on an iPad Pro tablet (screen size of 12.9 inches and a 2732×2048 resolution), using the Philips IntelliSpace Cognition application, which is approved by the US Food and Drug Administration as a Class II medical device (GUDID 00884838108554). Instructions and guidance were provided to the participants in both written and verbal form. The participants used verbal responses, touch of the keypad and drawing with a digital pencil or finger to perform the tests.

The research personnel were individually trained on the use of Philips IntelliSpace Cognition. Research personnel was always present during administration of the cognitive tests to ensure that the patients understood and followed instructions and to administer questionnaires and paper-and-pencil screeners. Research personal was instructed to provide minimal assistance with the cognitive tests. Most of the patients performed the tests independently, requiring very little supervision from the research personnel. The most frequently used guidance was prompting the patient to replay the test instructions. Each participant completed the assessment.

The predefined cognitive battery, included Rey’s Auditory Verbal Learning Test (RAVLT), Trail-Making Test (TMT) A and B, Clock Test Drawing and Copy, Star Cancellation Test, Rey-Osterrieth Complex Figure Test (ROCFT), Letter Fluency also known as the Controlled Word Association Test (COWAT), Digit Span Forward and Backward, and two additional cognitive tests; Category Fluency and the Symbol Number Matching Test (SNMT), also known as, or equivalent to, the Symbol Digit Modalities Test or Digit Symbol Substitution Test [13]. Digital versions of the Mini-Mental State Exam version 2, (MMSE) [14],, the Patient Health Questionnaire (PHQ-9) and General Anxiety Disorder-7 (GAD-7) were also administered. Only the MMSE, but not the two questionnaires are included in the analyses. The verbal responses were automatically transcribed by voice recognition software and the drawing tests were analyzed by proprietary computer vision algorithms. These automated annotation algorithms have been validated against human expert raters [4]. For participants who were unable to reach the last target on TMT (either A or B), and thus did not complete the test within time, the normed score was imputed based on the lowest norm percentile. This is in line with the TMT manual and prevents misrepresentation of a shorter duration due to participants not finishing the TMT. In the MCI cohort, this occurred 3 times for TMT-A and 12 times for TMT-B. In the CN cohort, this occurred twice for TMT-B.

### 2.3 Cognitive domain scores

We used a cognitive factor model to estimate a disease-agnostic profile in six cognitive domains: memory, executive functioning, processing speed, verbal processing, visual spatial processing, and working memory. These domain scores were derived from a structural equation model that was previously established in the healthy cohort referenced above [3]. Table 1 lists the neuropsychological tests and scores used to construct the cognitive domain scores (CDS). Note that cognitive domain scores are standard scores, adjusted for the effects of age, sex, and education.

**Table 1:** Descriptive statistics

|  |  | CN<br>(N = 83) | MCI<br>(N = 83) | p-value | effect<br>size | used in<br>CDS | used in<br>EMACC | used in<br>PACC |
| --- | --- | --- | --- | --- | --- | --- | --- | --- |
| Age (years) |  | 73.2 ± 7.12 | 74.2 ± 7.62 | 0.384 | -0.14 |  |  |  |
| Sex (n) | Female | 34 (41%) | 41 (49%) | 0.349 | 0.17 |  |  |  |
|  | Male | 49 (59%) | 42 (51%) |  |  |  |  |  |
| Education (years) |  | 17.8 ± 3.06 | 17.8 ± 3.07 | 0.960 | -0.02 |  |  |  |
| RAVLT (total score) | Learning Trials | 41.2 ± 7.08 | 30.7 ± 10.2 | < 0.001 | 1.21 | × | × | - |
|  | Immediate Recall | 7.51 ± 3.00 | 4.02 ± 3.27 | < 0.001 | 1.12 | × | - | - |
|  | Delayed Recall | 7.99 ± 3.04 | 4.71 ± 3.53 | < 0.001 | 1.00 | × | - | × |
| Trial Making (duration) | Test A | 45.4 ± 15.2 | 53.3 ± 28.3 | 0.030 | -0.35 | × | × | - |
|  | Test B | 116 ± 53.3 | 169 ± 133 | < 0.001 | -0.54 | × | × | - |
| Clock Test (total score) | Drawing | 18.4 ± 2.43 | 18.2 ± 3.28 | 0.674 | 0.07 | - | - | - |
|  | Copy | 19.6 ± 1.87 | 19.0 ± 3.31 | 0.162 | 0.22 | - | - | - |
| Star Cancellation (duration*) |  | 0.92 ± 0.25 | 1.15 ± 0.50 | < 0.001 | -0.61 | × | - | - |
| ROCFT (total score) | Copy | 30.0 ± 5.08 | 28.5 ± 7.20 | 0.126 | 0.24 | × | - | - |
|  | Immediate Recall | 9.88 ± 6.00 | 6.25 ± 5.74 | < 0.001 | 0.62 | × | - | - |
| Digit Span (total score) | Forward | 7.78 ± 2.04 | 6.36 ± 2.18 | < 0.001 | 0.68 | × | - | - |
|  | Backward | 6.14 ± 2.69 | 5.37 ± 2.37 | 0.052 | 0.31 | × | - | - |
| Letter Fluency (total score) |  | 40.0 ± 11.7 | 35.7 ± 11.3 | 0.015 | 0.38 | × | - | - |
| Category Fluency (total score) | Trial Animals | 20.4 ± 5.39 | 15.2 ± 5.05 | < 0.001 | 1.00 | - | × | × |
|  | Trial Vegetables | 12.3 ± 3.29 | 9.99 ± 4.07 | < 0.001 | 0.63 | - | - | - |
|  | Trial Fruit and Furniture | 12.6 ± 2.98 | 9.02 ± 3.09 | < 0.001 | 1.19 | - | × | × |
| SNMT (total score) |  | 37.8 ± 5.61 | 34.0 ± 8.63 | 0.027 | 0.46 | - | × | × |
| MMSE (total score) |  | 27.8 ± 1.40 | 26.5 ± 2.67 | < 0.001 | 0.61 | - | - | × |

CN = Cognitively Normal; MCI = Mild Cognitive Impairment; Effect Size is quantified by Cohen’s *d*, the *p*-values reflect a two-sample *t*-test or chi-square test; RAVLT = Rey’s Auditory Verbal Learning Test; duration\* = duration per correct cancellation in seconds; ROCFT = Rey-Osterrieth Complex Figure Test; SNMT = Symbol Number Matching Test; MMSE = Mini Mental State Examination version 2; The SNMT has a smaller number of observations for CN (*n* = 22). In the three columns on the right, the X marks indicate what neuropsychological tests/metrics are used for the Cognitive Domain Scores (CDS), Early AD/MCI Alzheimer’s Cognitive Composite (EMACC) or Preclinical Alzheimer’s Cognitive Composite (PACC).

### 2.4 Disease-specific cognitive composites

We evaluated two disease-specific composite scores: the Early AD/MCI Alzheimer’s Cognitive Composite (EMACC) and the Preclinical Alzheimer’s Cognitive Composite (PACC). EMACC was specifically designed as a cognitive endpoint for clinical trials of early AD [9]. It includes neuropsychological tests from the domains of memory, executive functioning, and processing speed. The exact neuropsychological tests, or versions of them, used to construct the EMACC vary slightly between studies [9, 8]. In this study, EMACC was defined similar to the Mayo Clinic Study of Aging, using the RAVLT learning total score, category fluency-animals and category fluency fruit and Furniture, TMT-A, TMT-B, and SNMT.

The PACC is designed for the detection of subtle cognitive changes in the early stages of the disease continuum [10, 11], and includes neuropsychological tests from the domains of memory and executive functioning, as well as a measure of general cognition. In this study, the PACC was defined similar to the extended definition using a measure of semantic processing [15]. As in the EMACC, the exact tests used to construct the composite score vary slightly between various studies and clinical trials [10, 12, 16, 17, 15, 18]. Instead of the free and cued selective reminding test (FCSRT), we used the RAVLT delayed recall, akin to the PACC definitions that use the California Verbal Learning Test delayed recall [10, 16]. Table 1) lists the neuropsychological tests/scores used for the EMACC and PACC. Both EMACC and PACC use z-scoring to normalize the test scores in order to combine them into a single composite score. We estimate the z-scores using the same normative cohort used for the cognitive domain scores. Note that the z-transform does not correct the disease-specific composites for age, sex, or education.

### 2.5 Statistics

Statistical analysis was performed with R v4.1.1 and the base R stats package [19]. This includes t-tests, logistic regression models, and Cohen’s d. The figures and tables were generated with ggplot2 v3.3.3 [20] and the table1 package v1.4.2 [21]. The receiver operating characteristic (ROC) curves were calculated with the pROC package v1.18.0 [22] and statistically compared using bootstrapping tests for two correlated ROC curves (n=2000). Evaluation metrics for sensitivity, specificity, and area under the curve (AUC), were calculated with the cutpointr package v1.1.1 [23]. The product of sensitivity by specificity was used to determine an optimal position on the ROC curve.

The cohort of cognitively normal (CN) older adults was selected with a non-parametric matching procedure [24] from a normative cohort [4, 3]. First, we removed all CNs with an MMSE < 25 and RAVLT immediate recall score one standard deviation below the norm, similar to ADNI criteria for healthy controls [12]. Next, we employed nearest-neighbor matching based on a propensity score, as implemented in the MatchIt package v 4.3.2 [25]. The propensity score was estimated using a logistic regression model with age, sex, and education level. Table 1 shows the descriptive statistics that compare the CNs matched with the MCI patients. The p-values are based on two-sample t-tests, or in the case of sex, a chi-square test. The matching procedure ensures that comparisons of various tests, cognitive domain scores, and disease-specific composites between CN and MCI are not simply driven by demographic factors.

## 3 Results

### 3.1 Descriptive results

The cohort of MCI patients performed significantly worse on all tests (see Table 1), with the exception of the ROCFT Copy and the Clock Drawing Test, where the performance differences were not significant. Based on the effect size, quantified by Cohen’s d, the RAVLT Learning Trials showed the largest difference between CN and MCI, and similar effects were observed with RAVLT Immediate Recall, Delayed Recall, and Category Fluency. The effect size was much smaller for the other tests. The variance was greater in the MCI cohort for each test, except Digit span backward and Category Fluency (Animals).

The cognitive domain scores were ranked by their relative effect size. MMSE (Cohen’s d = 0.61) was used as a baseline (Figure 1). The memory domain showed the largest effect size (Cohen’s d = 1.24), followed by executive functioning (Cohen’s d = 0.99), visual spatial processing (Cohen’s d = 0.83), working memory (Cohen’s d = 0.79) and processing speed (Cohen’s d = 0.66) – all greater than MMSE. Verbal processing showed the smallest effect size (Cohen’s d = 0.40), consistent with the observation that typical MCI patients might decline, but commonly show no clinical impairment in verbal processing [26].

**Figure 1:**
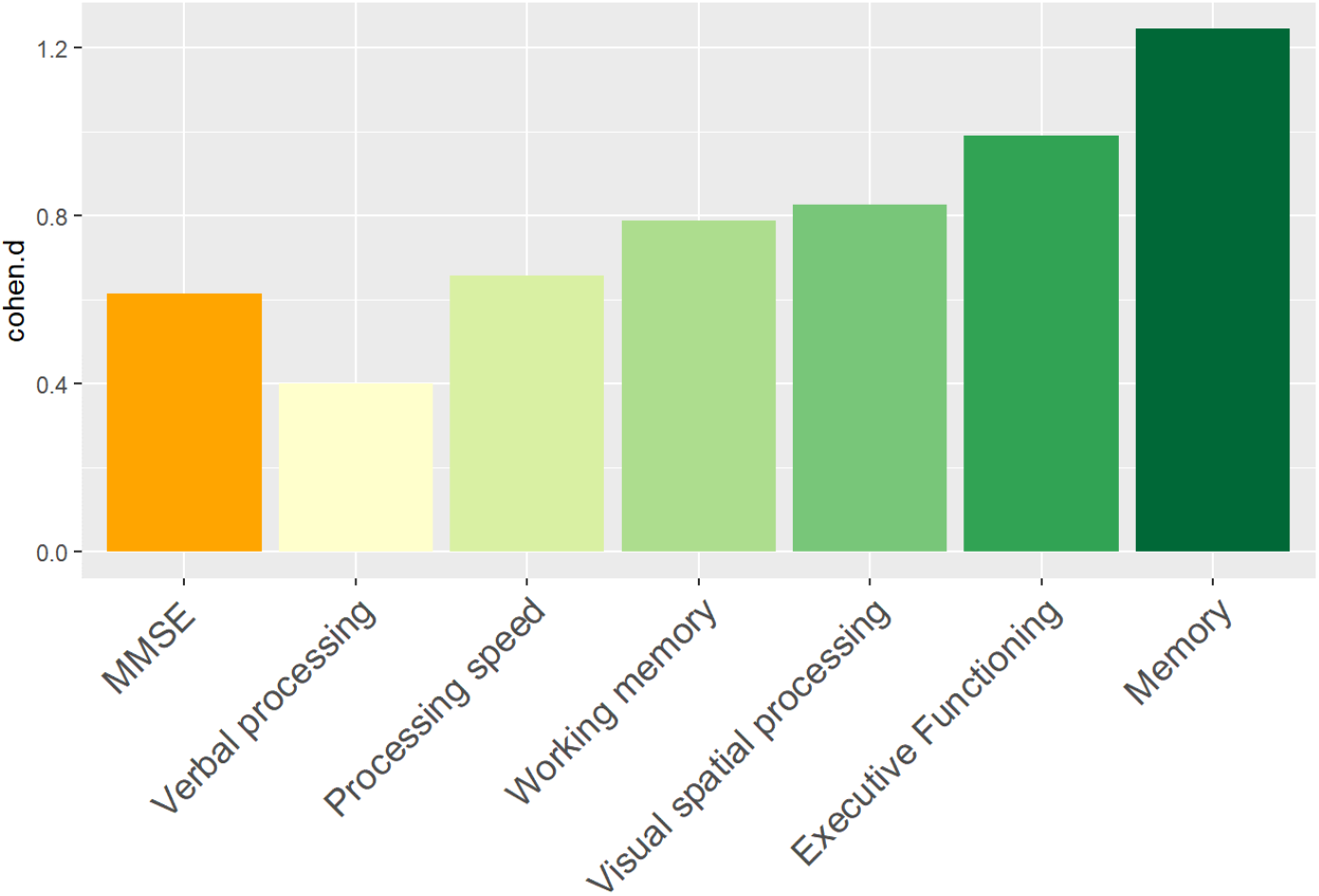
Effect size for MMSE and cognitive domain scores.

### 3.2 Sensitivity and specificity

We used logistic regression models to classify the diagnosis of CN versus MCI based on demographic factors, MMSE, cognitive domain scores, and disease-specific composites. We calculated sensitivity, specificity, and area under the curve (AUC) as shown in Table 2. We also visualized the ROC curves for a selection of models (Figure 2). The prevalence estimate (and majority baseline) is 0.5 since the CN and MCI groups are balanced. The AUC of models that use age, sex, and education is at the chance level as a consequence of the matching procedure. Consistent with the largest effect size (Figure 1), we found that of the cognitive domains memory and executive functioning were the strongest predictors of the diagnosis, while verbal processing performed slightly below the MMSE. We also examined whether a combination of cognitive domain scores would improve classification by running logistic regression models for all possible combinations of two domain scores, including an interaction term. The logistic regression model with memory * executive functioning performed best and is included in Table 2. However, it only marginally improved AUC relative to a model with only memory (see also statistical comparisons below). The model that used the PACC showed the highest AUC, closely followed by the EMACC.

**Table 2:** Sensitivity, Specificity and AUC

| Predictor | Sensitivity | Specificity | AUC | CI 95% |
| --- | --- | --- | --- | --- |
| PACC | 0.87 | 0.77 | 0.88 | 0.83-0.93 |
| EMACC | 0.82 | 0.82 | 0.87 | 0.81-0.92 |
| Memory * Executive Functioning | 0.86 | 0.71 | 0.83 | 0.77-0.89 |
| Memory | 0.90 | 0.61 | 0.80 | 0.74-0.88 |
| Executive Functioning | 0.78 | 0.58 | 0.75 | 0.68-0.82 |
| MMSE (reference) | 0.67 | 0.57 | 0.62 |  |
| Verbal Processing | 0.66 | 0.53 | 0.61 | 0.52-0.69 |
| Age | 0.73 | 0.37 | 0.54 | 0.45-0.63 |
| Education | 0.51 | 0.54 | 0.51 | 0.42-0.59 |
| Sex | 0.41 | 0.51 | 0.46 | 0.38-0.53 |

*PACC = Preclinical Alzheimer’s Cognitive Composite; EMACC = Early AD/MCI Alzheimer’s Cognitive Composite, MMSE = Mini Mental State Examination, AUC = Area Under the Curve, CI = Confidence Interval*

**Figure 2:**
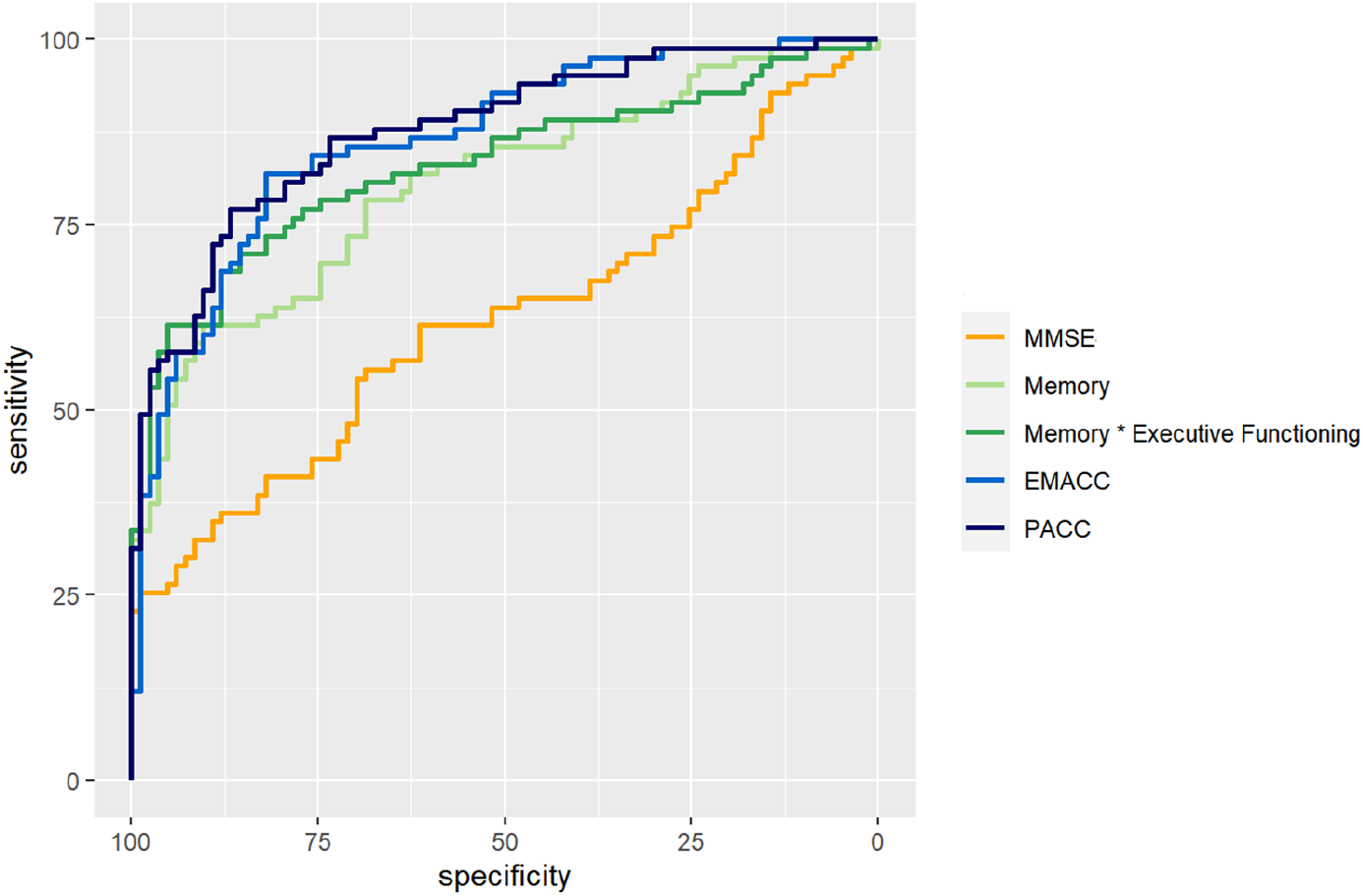
ROC curves

The results of pairwise statistical comparisons for a selection of ROC curves can be seen in 2). First, we compared the ROC curves with the ROC curve for MMSE, Memory > MMSE (D = 3.88; p < 0.001), Memory * Executive Functioning > MMSE (D = 4.48; p < 0.001), EMACC > MMSE (D = 5.39; p < 0.001) and PACC > MMSE (D = 6.64; p < 0.001). In the comparisons with the Memory domain score, we found smaller differences: Memory * Executive Functioning > Memory (D = 1.68; p = 0.047), EMACC > Memory (D = 1.63; p = 0.052), PACC > Memory (D = 1.98; p = 0.024). When comparing the disease-specific cognitive composites to Memory * Executive Functioning, we found no statistical differences: EMACC > Memory * Executive Functioning (D = 0.85; p = 0.197) and PACC > Memory * Executive Functioning (D = 1.20; p = 0.116). Similarly, we did not find any evidence for a difference between the PACC > EMACC (D = 0.553; p = 0.290).

## 4 Discussion

Our study demonstrates the clinical validity of a digital cognitive platform, IntelliSpace Cognition, in patients with MCI. We found that performance in MCI patients was significantly worse compared to cognitively normal adults, on almost all neuropsychological tests. Furthermore, performance on some tests was much more impaired than on others. Specifically, from the six cognitive domains investigated, the memory domain was the most impaired. Finally, AD-specific composite scores showed greater sensitivity and specificity than the individual cognitive domain scores, individual neuropsychological tests, and the MMSE.

When comparing the performance of individual neuropsychological tests between CN and MCI (see Table 1), RAVLT, a test of learning and memory, was most affected in MCI, consistent with diagnostic criteria [5]. Category fluency showed the second largest effect size, while letter fluency was much less impaired. This replicates previous findings [27, 28, 29]. The cognitive domain score reflects these individual test results: the memory domain demonstrated the greatest impairment in the MCI group, which follows from factor model dependencies [4]. Executive functioning was the second most impaired domain in MCI; a finding that also replicates previous work [30, 31, 32]. and may be related to the presence of vascular-dominant cases. The executive functioning domain includes the TMT-A and TMT-B, Star Cancellation Test, ROCFT Delayed Recall, Letter Fluency and Forward and Backward Digit Span, but not RAVLT. We also compared logistic regression models based on any combination of two domain scores. The combination of memory by executive functioning showed the highest AUC (see Table 2). This model slightly outperformed a logistic classifier that used only the memory domain. This result is consistent with current perspectives on cognitive domains in AD and on how to construct cognitive composites for clinical trials [7, 33]. Together, these results confirm the recommendation to use neuropsychological tests in the domain of memory and executive functioning in patients with MCI.

We constructed two digital cognitive composite scores: the EMACC and PACC[10, 9]. The EMACC and PACC classified CN versus MCI patients with an AUC of .87 and .88 respectively (Table 2). Both AD-specific composites outperform the MMSE, individual neuropsychological tests, and individual cognitive domain scores. The logistic regression model that included both memory, executive functioning and an interaction term had an AUC of 0.83. This model was not statically different from the EMACC and PACC. These findings are consistent with the notion that MCI should not be defined by impaired cognition in a single domain. It is worth noting that the PACC includes the MMSE, as a measure of global cognition; in contrast, the EMACC does not. Our findings do not elucidate if a measure of global cognition improves detection, as the difference between PACC and EMACC was also not significant. Both composites were originally designed to quantify early cognitive impairment. Therefore, it is no surprise that they are sensitive to differences between the CN and MCI patients. Importantly, these composites were designed with conventional paper-and-pencil neuropsychological tests. Here, we replicate this work, but using a digital cognitive assessment platform.

### 4.1 Limitations

First, this was a single-center study with a modest sample size of 83 MCI patients. A larger sample size would provide stronger statistical power to detect subtle differences between the various tests. Second, no amyloid-*β* or tau biomarkers were obtained. Therefore, we cannot characterize these patients in terms of AD research stages [34, 35]. The inclusion criteria were based on current clinical guidelines and included MRI. The resulting MCI cohort was probably heterogeneous, consisting of patients with and without amyloid, tau and some degree of vascular pathology. This might as well be regarded as a strength, as it mirrors the current diagnostic pathway. Nevertheless, recent biomarker-based studies reported similar results when using the PACC composite score [36, 2]. Third, traditional paper-and-pencil tests were not administered in patients with MCI. This would have required a counterbalanced design and was beyond the scope of this study. Hence, we cannot determine if the sensitivity or specificity of our digital cognitive platform is smaller or greater than traditionally administered tests with manual scoring. However, test scores are similar to scores reported for traditional paper-and-pencil tests in MCI, and the equivalence of IntelliSpace Cognition to paper-and-pencil tests has previously been demonstrated in CN adults [3].Thus, although we cannot exclude minor differences, we can infer that the platform performs on-par with traditional paper-and-pencil tests.

### 4.2 Future directions

The IntelliSpace Cognition platform is disease agnostic and includes a range of neuropsychological tests that are clinically used in multiple disorders, including Alzheimer’s disease, Parkinson’s disease, frontotemporal dementia, traumatic brain injury, stroke and multiple sclerosis. The digital battery is therefore ideally suited to gather high-quality data in a clinical care setting. This can enable research into cognitive phenotypes within, or across, heterogeneous diseases. Since, IntelliSpace Cognition is approved by the FDA, the battery is well positioned to establish the cognitive baseline for large, clinically heterogeneous populations, which can be the basis for different investigations on early diagnosis, outcome prediction, or treatment response.

One question remains: How important will detailed cognitive profiles remain, as the field moves towards more biological definitions of dementia? It is evident, that a decision to start treatment with amyloid-or tau antibodies, will require confirmation of the underlying pathology. Furthermore, recent work suggests that cognitively normal adults with a positive amyloid and tau biomarker status, are destined to decline [37]. Yet, many patients do not fit this profile and the boundaries between positive and negative biomarker status are often not clear cut. For example, patients that only harbor tau, also exhibit impairments in different cognitive domains [38]. Thus, it is likely that cognitive measurements will continue to improve a clinical forecast. An accurate assessment of cognition, together with a the assessment of pathology, is crucial for a decision to start treatment. Digital tools will have an advantage over paper and pencil methods for operational reasons, and are likely to play a key role in treatment decision and monitoring of treatment efficacy [39].

In conclusion, the IntelliSpace Cognition platform can be used to characterize cognitive deficits in patients with MCI. Compared to other digital assessment tools, the IntelliSpace Cognition battery is more comprehensive. It hosts digital neuropsychological tests that allow constructions of cognitive profiles across neurological diseases. This can be used to create a personalized, disease-specific assessment, utilizing standard metrics, as well as new digital data points. Further research should focus on other diseases as well as on combining cognitive data with Alzheimer’s disease biomarkers.

## Data Availability

Most data produced in the present work are contained in the manuscript. Additional information can be requested at https://www.usa.philips.com/healthcare/solutions/neurology/digital-cognitive-assessment

## 5 Author Contributions

Study design: NP, GvE, MS, ESW, BS, SH

Data acquisition: NP, ESW, RPD, BFA, SH, MDY

Data analysis: WH, GvE, MC

Interpretation: WH, GvE, BS, NP

Manuscript: WH, GvE, MS, MC, BS, BA, MY, SH, RPD, BA, ESW, NP

## 6 Acknowledgements

The authors thank Dr. Murray Gillies, former head of the ISC venture, for his support. Dr. Christopher Gantz from the Sidney Kimmel Cancer Center provided valuable help with recruitment practices. We also thank Dr. Laura Klaming for her feedback on the manuscript

